# Unspecific *post-mortem* findings despite multiorgan viral spread in COVID-19 patients

**DOI:** 10.1101/2020.05.27.20114363

**Authors:** Myriam Remmelink, Ricardo De Mendonça, Nicky D’Haene, Sarah De Clercq, Camille Verocq, Laetitia Lebrun, Philomène Lavis, Marie-Lucie Racu, Anne-Laure Trépant, Calliope Maris, Sandrine Rorive, Jean-Christophe Goffard, Olivier Dewitte, Lorenzo Peluso, Jean-Louis Vincent, Christine Decaestecker, Fabio Silvio Taccone, Isabelle Salmon

## Abstract

**Background:** *Post-mortem* studies can provide important information for understanding new diseases and small autopsy case series have already reported different findings in COVID-19 patients.

**Methods:** We evaluated whether some specific *post-mortem* features are observed in these patients and if these changes are related to the presence of the virus in different organs. Complete macroscopic and microscopic autopsies were performed on different organs in 17 COVID-19 non-survivors. Presence of SARS-CoV-2 was evaluated with immunohistochemistry (IHC) in lung samples and with real-time reverse-transcription polymerase chain reaction (RT-PCR) test in lung and other organs.

**Results:** Pulmonary findings revealed early-stage diffuse alveolar damage (DAD) in 15 out of 17 patients and microthrombi in small lung arteries in 11 patients. Late-stage DAD, atypical pneumocytes and/or acute pneumonia were also observed. Four lung infarcts, two acute myocardial infarctions and one ischemic enteritis were observed. There was no evidence of myocarditis, hepatitis or encephalitis. Kidney evaluation revealed the presence of hemosiderin in tubules or pigmented casts in most patients. Spongiosis and vascular congestion were the most frequently encountered brain lesions. No specific SARS-CoV-2 lesions were observed in any organ. IHC revealed positive cells with a heterogeneous distribution in the lungs of 11 of the 17 (65%) patients; RT-PCR yielded a wide distribution of SARS-CoV-2 in different tissues, with 8 patients showing viral presence in all tested organs (i.e. lung, heart, spleen, liver, colon, kidney and brain).

**Conclusions:** In conclusion, autopsies revealed a great heterogeneity of COVID-19-related organ injury and the remarkable absence of any specific viral lesions, even when RT-PCR identified the presence of the virus in many organs.

## Background

Coronaviruses, including severe acute respiratory syndrome coronavirus (SARS-CoV) and Middle East respiratory syndrome coronavirus (MERS-CoV), cause severe acute respiratory failure, which is associated with high mortality rates (1). The novel SARS-CoV-2 strain exhibits phylogenetic similarities to SARS-CoV and causes coronavirus disease 2019 (COVID-19), which has caused more than 280,000 deaths worldwide so far. As the pandemic has progressed, the pathophysiology of this viral infection has become clearer; in particular, it has been shown that SARS-CoV-2 can directly alter cell function by a link to the angiotensin converting enzyme 2 (ACE2) receptor, which is almost ubiquitous in the human body (2).

Nevertheless, the mechanisms behind the high mortality and severe organ dysfunction associated with COVID-19 remain poorly understood. Controversies exist regarding the occurrence of fatal complications, such as pulmonary embolism or diffuse endothelial injury (3, 4), as well as on the roles of direct viral cellular injury or concomitant comorbidities in the fatality of this disease (5).

In this setting, autopsy is of great importance to help physicians understand the biological characteristics and the pathogenesis of COVID-19. Most of the previously reported *post-mortem* findings focused on lung morphology and few data are available on complete *post-mortem* analyses of other organs (6, 7). The aim of this study was therefore to investigate the presence of specific features of viral injury as well as the distribution of the virus in different organs of patients who died from COVID-19.

## Methods

### Study design

In this *post-mortem* study, we included the first 17 adult patients (>18 years) who died in our hospital (either in a COVID-19 unit or an intensive care unit) from March 13, 2020 with confirmed SARS-CoV-2 infection (i.e. positive RT-PCR assay on nasopharyngeal swab and/or broncho-alveolar lavage specimen). Autopsies were performed 72 to 96 hours after death to ensure the safety of the autopsy team. Exclusion criteria were lack of family consent and a delay of more than five days after death. The study protocol was approved by the local ethics committee (P2020/218).

### Data collection

We collected demographics, comorbidities, relevant clinical data, the results of chest computed tomography scan, and if available, microbiological tests and medical treatments (e.g. hydroxychloroquine, antivirals or antibiotics, and use of organ support). Acute respiratory distress syndrome (ARDS) and acute kidney injury (AKI) were defined according to standard definitions (8, 9).

### Post-mortem procedure

The Belgian Sciensano guidelines were integrated into our *post-mortem* procedure (10). Personal protective equipment consisted of two superposed disposable latex gloves, plastic sleeves, FFP3 mask, scrub hat, clear face visor, surgical gown plus plastic apron, rubber boots. In the *post-mortem* room, dirty and clean circulations were used in the airlocks to allow decontamination. For 11 cases, brain samples were obtained using a safety procedure with drills and protective devices to avoid air dispersion, as described in the Additional file 1 (Additional Material).

Using standard surgical pathology processing, complete sets of tissue samples were collected for diagnosis and biobanking. The material was biobanked by Biobanque Hôpital Erasme-ULB (BE_BERA1), CUB Hôpital Erasme; BBMRI-ERIC. The banked material consists of 6 samples per organ, including trachea, thyroid, lymph nodes, heart, spleen, bone marrow, kidney, bladder, liver, stomach, colon, and brain. For the lungs, we collected six samples per lobe (i.e. a total of 30 samples), except for two patients who had undergone lobectomy for cancer and from whom only 18 samples were taken. For safety reasons, complete brain removal was not allowed, but, with the help of a neurosurgeon, we developed a new procedure to obtain between 12 and 51 samples from different brain regions, as detailed in the Additional file 1 (Additional Material). Formalin-fixed paraffin-embedded (FFPE) tissues underwent standard processing to provide hematoxylin and eosin (H&E)-stained sections. Special stains and immunohistochemistry (IHC) were used for lung (Masson’s trichrome, periodic acid-Schiff [PAS], Gomori-Grocott, anti-CMV IHC, anti-HSV IHC, anti-Pneumocystis J IHC) and kidney (PAS, Masson’s trichrome, Jones methenamine silver) samples.

### Morphological analysis

Morphological analysis was performed on H&E stained glass slides using the SecundOs digital platform (TribVn Health Care, Chatillon, France) for digital diagnosis, after acquisition of whole slide digital scans (40x magnification) using a Nanozoomer 2.0 HT slide scanner (Hamamatsu, Hamamatsu City, Japan).

### SARS-CoV-2 detection by immunohistochemistry

Since no antibody against SARS-CoV-2 has been validated for IHC on FFPE tissues, we selected an anti-SARS-nucleocapsid protein antibody. Standard IHC was applied as previously described to 4-μm-thick *post-mortem* lung sections (one sample for each lung lobe per patient) to display SARS-nucleocapsid protein (Invitrogen, PA1-41098, dilution 1:50) on Dako Omnis (Agilent Technologies, Santa Clara, CA, USA) using the Envision Flex detection system according to the manufacturer’s protocol (11). The sections were counterstained with hematoxylin. Negative tissue controls were obtained from patients who had an autopsy before the COVID-19 pandemic. Semi-quantitative IHC evaluation was performed by two senior pathologists (ND, MR) as follows: negative (−); between one and five positive cells per whole slide (scattered cells, +); more than five cells per whole slide but no foci (isolated cells, ++); and with foci (more than 10 cells in one 20X field, +++).

### SARS-CoV-2 detection by rRT-PCR

Total nucleic acid was extracted from FFPE tissues using the Maxwell RSC DNA FFPE Kit (reference: AS1450. Promega Corporation, Madison, WI, USA) and the Promega Maxwell extractor, following the protocol described by the manufacturer. One-step RT-PCR assays specific for the amplification of SARS-CoV-2 E envelope protein gene were adapted from a published protocol (12). Briefly, 4 μL of RNA (100 ng) was amplified in 20 μL reaction mixture containing 5 μL of TaqMan Fast Virus 1-step master mix (Life Technologies), 0.4 μM of each forward (ACAGGTACGTTAATAGTTAATAGCGT) and reverse (ATATTGCAGCAGTACGCACACA) primers and 0.2 μM of probe (FAM-ACACTAGCCATCCTTACTGCGCTTCG-BBQ). The amplification condition was 50°C for 10 min for reverse transcription, followed by 95°C for 20 seconds and then 45 cycles of 95°C for 3 seconds and 58°C for 30 seconds. A clinical sample highly positive for SARS-CoV-2 was diluted 1:1000 and used as a positive control in each analysis. A clinical sample obtained from a patient who was autopsied before the COVID-19 pandemic was used as a negative control. The quality of the RNA from the samples showing negative results was assessed by amplification of the human *MET* RNA according to a validated ISO:15189 accredited method used as a routine diagnostic method in our laboratory.

### Statistical analysis

Data are reported as counts (percentage) or medians [interquartile ranges (IQRs)]. All data were analyzed using GraphPad Prism Version 8.4.2 (GraphPad Software, San Diego, CA, USA).

## Results

### Study cohort

The main characteristics of the study cohort (12 males out of 17; median age 72 [62-77] years) are given in Table 1. The median time from admission to death was 13 [8-15] days. All except two patients had at least one comorbidity, including hypertension (n=10), diabetes (n=9), cerebrovascular disease (n=4), coronary artery disease (n=4) and solid cancer (n=4). None of the patients had tested positive on admission for the Respiratory Syncytial Virus or influenza A and B viruses. Eleven patients died in the ICU and 6 on the medical ward; the main causes of death were respiratory failure (n=9) and multiple organ failure (n=7). Laboratory data are reported in Additional file 2 (Table S1).

**Table 1:**
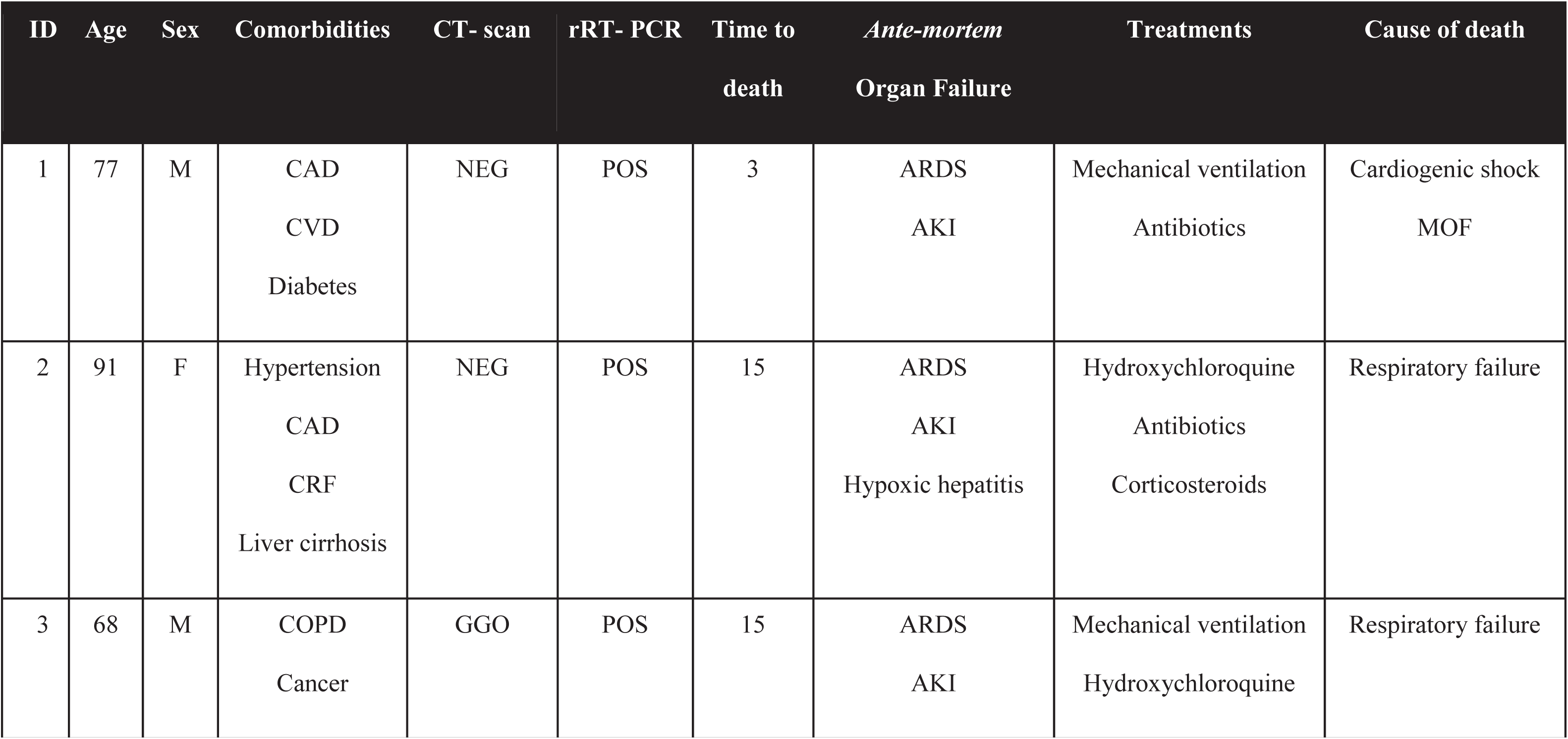
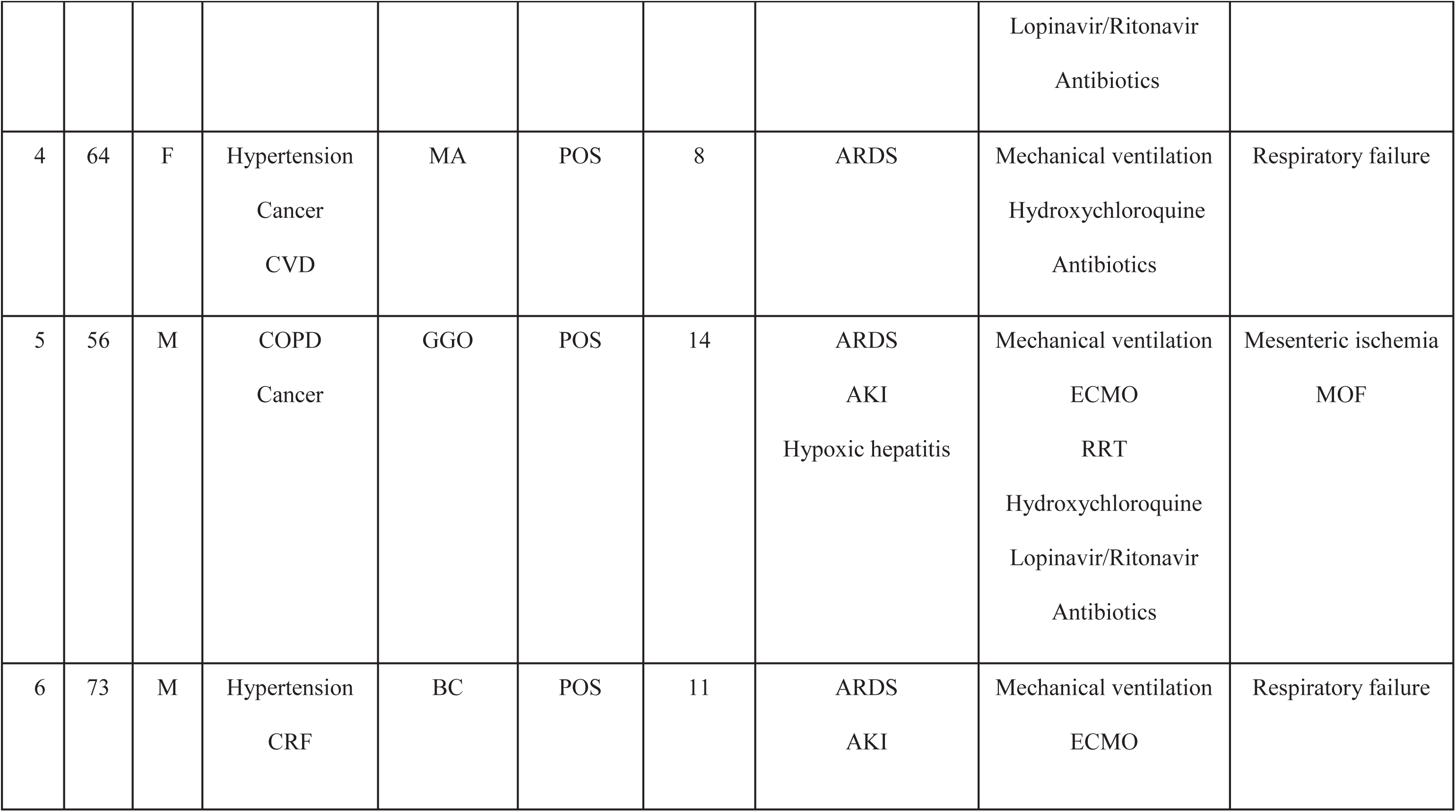
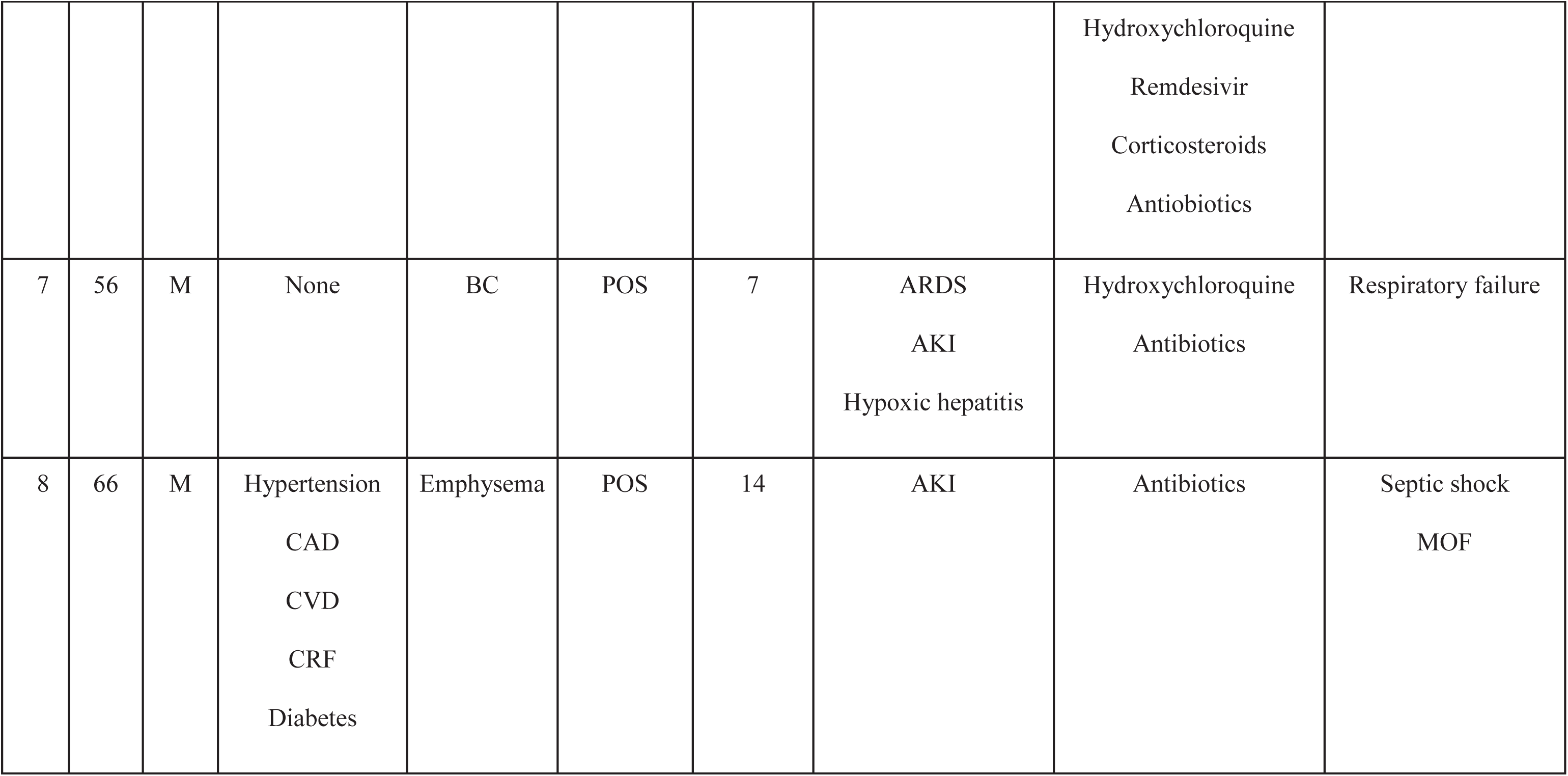
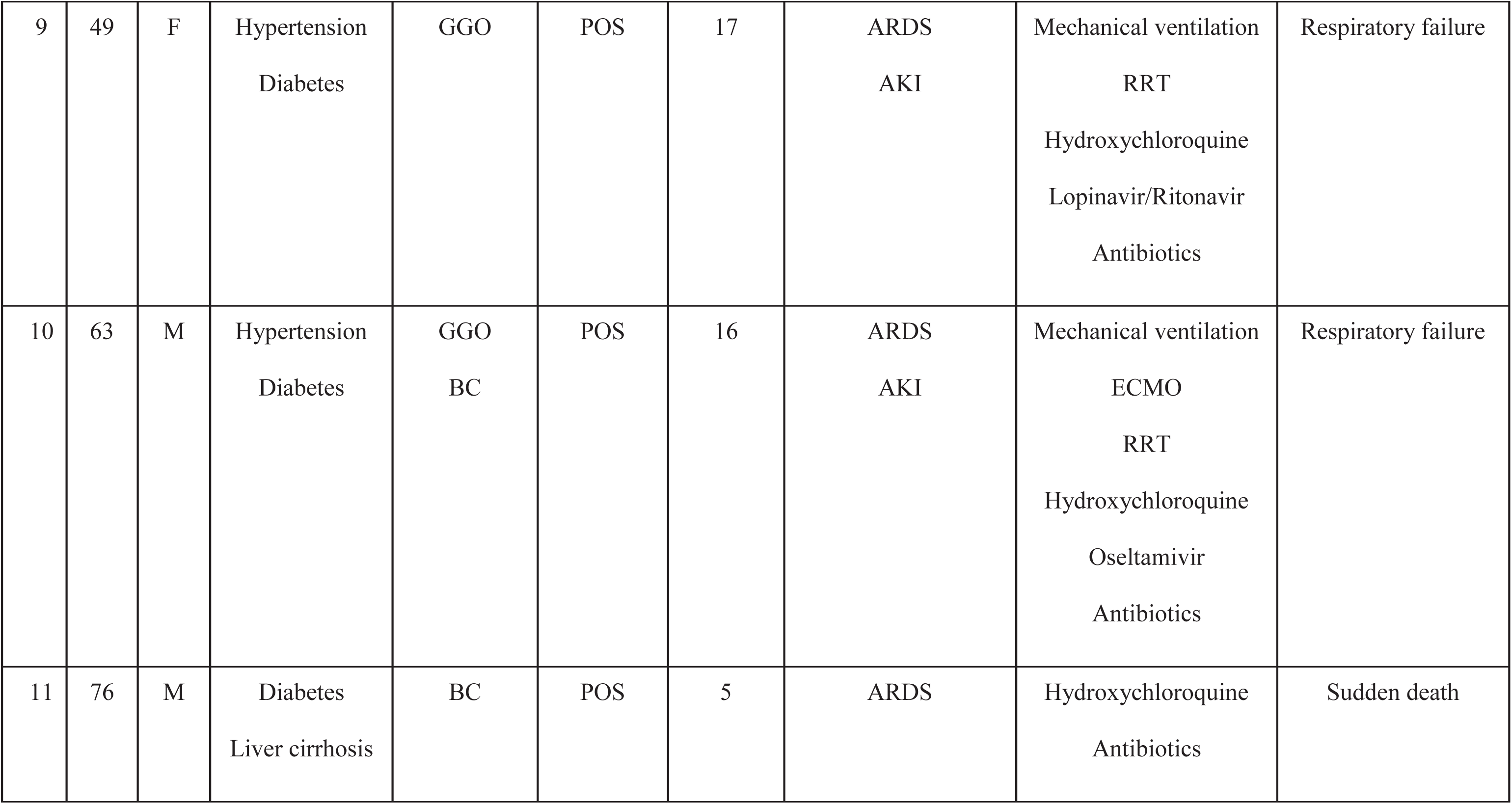
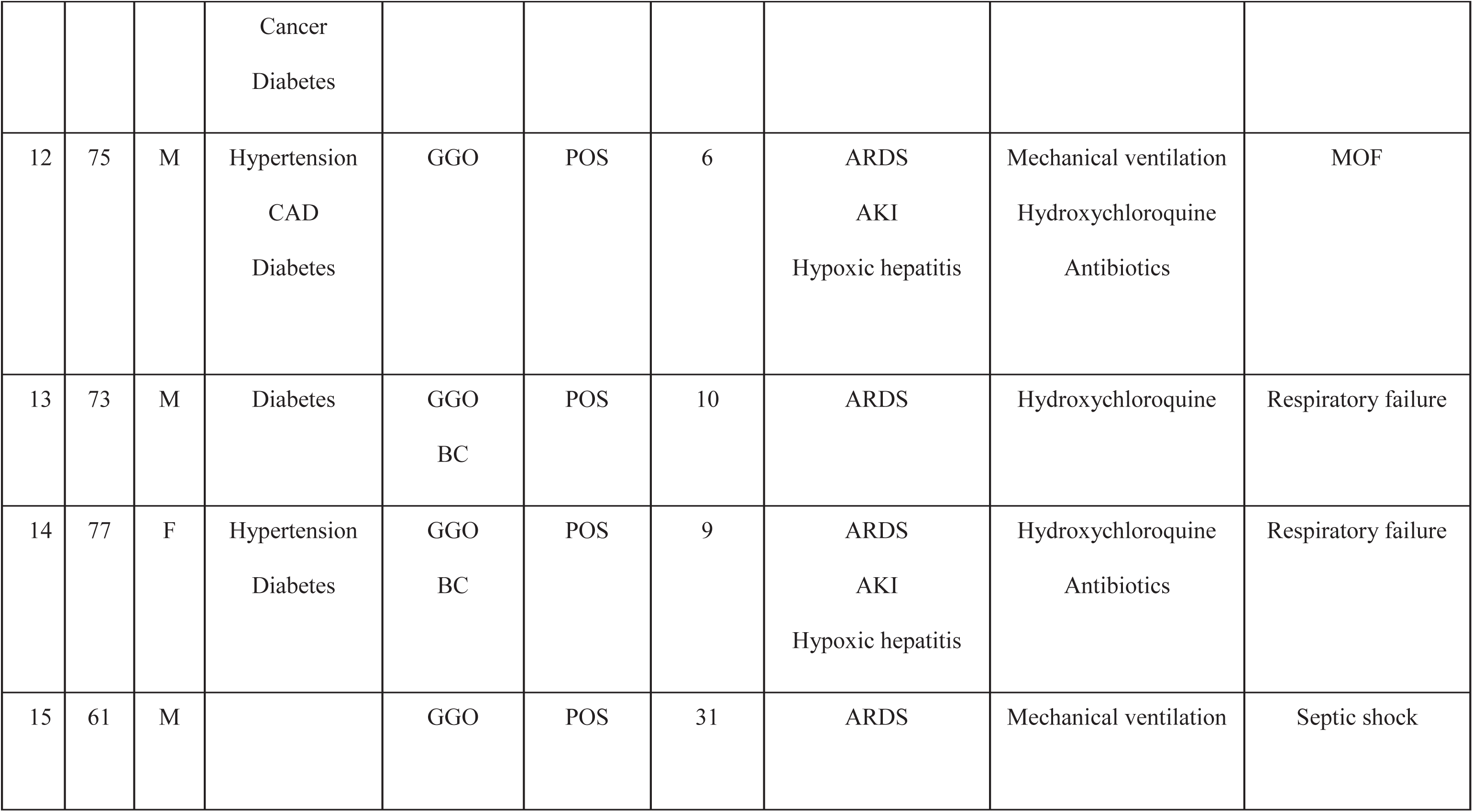
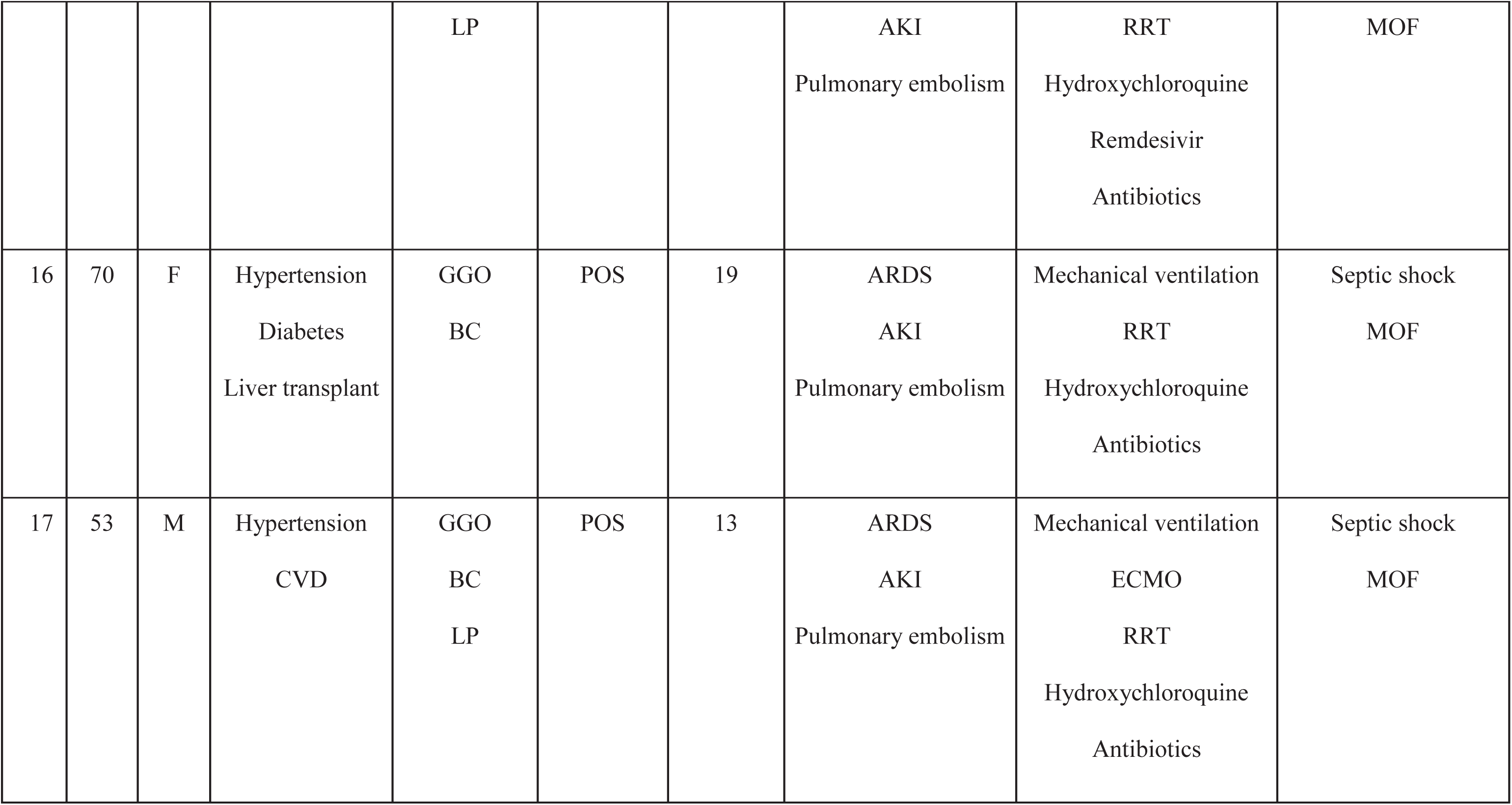
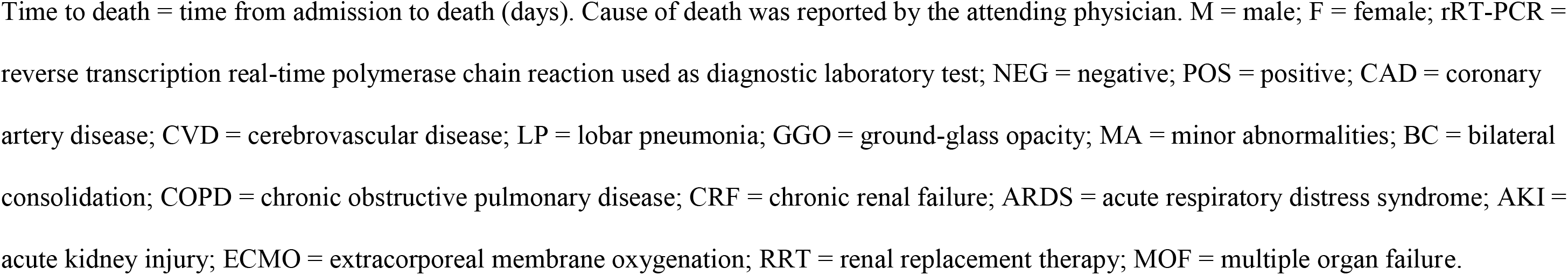
Characteristics of the study population.

### Macroscopic Findings

One patient had had a left pneumonectomy and one patient a right bilobectomy. The lungs were typically heavy and the lung parenchyma had a diffuse firm consistency with red/tan and patchy dark/red areas of hemorrhage. Thrombi were found in the large pulmonary arteries in 2 patients and lung infarction in 4 patients. Pleural adhesions associated with pleural effusions were observed in 4 cases. We observed cardiomegaly in 14 and and hepatomegaly in 5 patients. The kidneys were often enlarged, with a pale cortex and petechial aspect but no hemorrhage or infarct. The gut had advanced *post-mortem* autolysis with no evidence of specific lesions, except for one patient who had ischemic enteritis. In the 11 patients for whom brain samples were available, one had had a recently drained subdural hematoma and another had a cerebral hemorrhage.

### Microscopic Findings

As shown in Figures 1 and 2 and Additional file 3 (Table S2), the main pulmonary findings included early-stage diffuse alveolar damage (DAD), which consisted of interstitial and intra-alveolar edema, with variable amounts of hemorrhage and fibrin deposition, hyaline membranes, minimal interstitial mononuclear inflammatory infiltrate and type II pneumocyte hyperplasia. Microthrombi were noted in the small pulmonary arteries in 11 patients. Ten of the 17 patients also had advanced DAD lesions (i.e. fibroblastic proliferation within the interstitium and in the alveolar spaces); 8 patients had evidence of acute pneumonia or broncho-pneumonia, 4 had atypical pneumocytes and three syncytial multinucleated giant cells. We observed no viral inclusions or squamous metaplasia.

**Figure 1:**
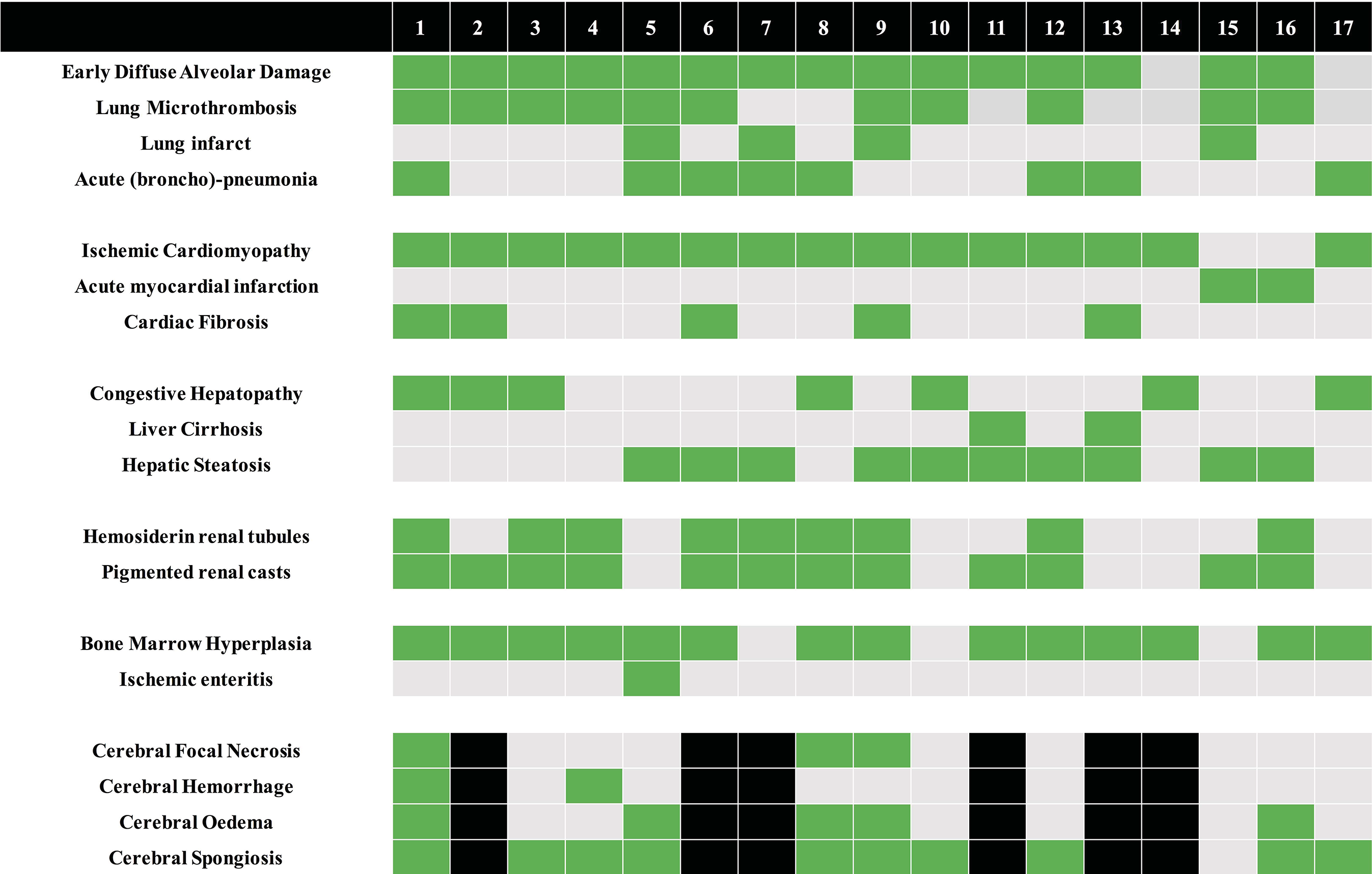
Main histological findings. Green = finding present; Gray = finding absent; Black = unavailable.

**Figure 2:**
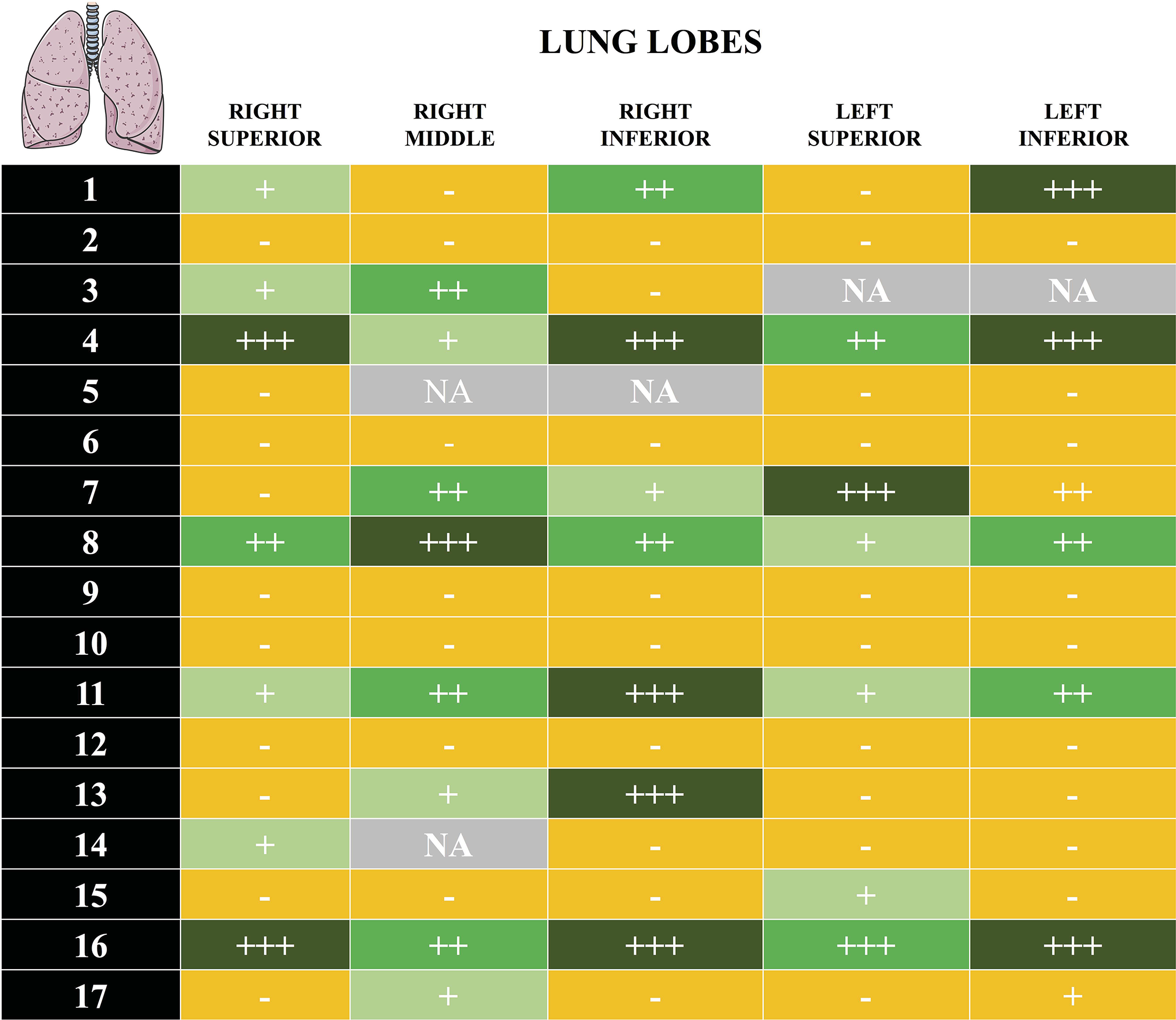
Pulmonary histological findings. A) Early stage diffuse alveolar damage (DAD): hyaline membrane (H&E, 50x magnification) with a zoom on a giant cell (100x magnification). B) Fibrin thrombi in a pulmonary artery (H&E, 50x magnification). C) Late stage DAD: fibroblastic proliferation (H&E, 50x magnification). D) Late stage DAD: fibroblastic proliferation (Trichrome staining, 50x magnification). E) Acute pneumonia (H&E, 50x magnification). F) Anti-SARS-CoV immunohistochemistry (IHC)-positive cells (200x magnification)

Fifteen patients had signs of chronic ischemic cardiomyopathy of different severities and 2 patients had signs of acute myocardial infarction; there was no evidence of contraction bands or myocarditis. Histological evaluation of the kidneys was limited because of moderate to severe *post-mortem* autolysis; occasional hemosiderin granules were observed in the tubular epithelium in 9 patients and pigmented casts in 12. In the medulla, edematous expansion of the interstitial space without significant inflammation was observed in 4 patients. Chronic renal lesions (i.e. nodular mesangial expansion and arteriolar hyalinosis, glomerulosclerosis or chronic pyelonephritis) were also observed; no microthrombi were identified, but one patient had a thrombus in an interlobar artery.

Liver examination revealed congestive hepatopathy and steatosis, but no patchy necrosis, hepatitis or lobular lymphocytic infiltrate. The histological changes in the abdominal organs including the esophagus, stomach and colon are reported in Additional file 3 (Table S2); most of the findings were related to chronic underlying diseases, except for one case of ischemic enteritis.

Brain samples showed cerebral hemorrhage or hemorrhagic suffusion (n=8), focal ischemic necrosis (n=3), edema and/or vascular congestion (n=5), and diffuse or focal spongiosis (n=10). We found no evidence of viral encephalitis or vasculitis, isolated neuronal necrosis or perivascular lymphocytic infiltration.

### SARS-CoV-2 detection in the lungs by IHC

SARS-CoV-2 was identified by IHC in the lungs of 11 of the 17 patients (Figure 3). However, there was large variability in the distribution of SARS-CoV-2 positive cells in the lung parenchyma.

**Figure 3:**
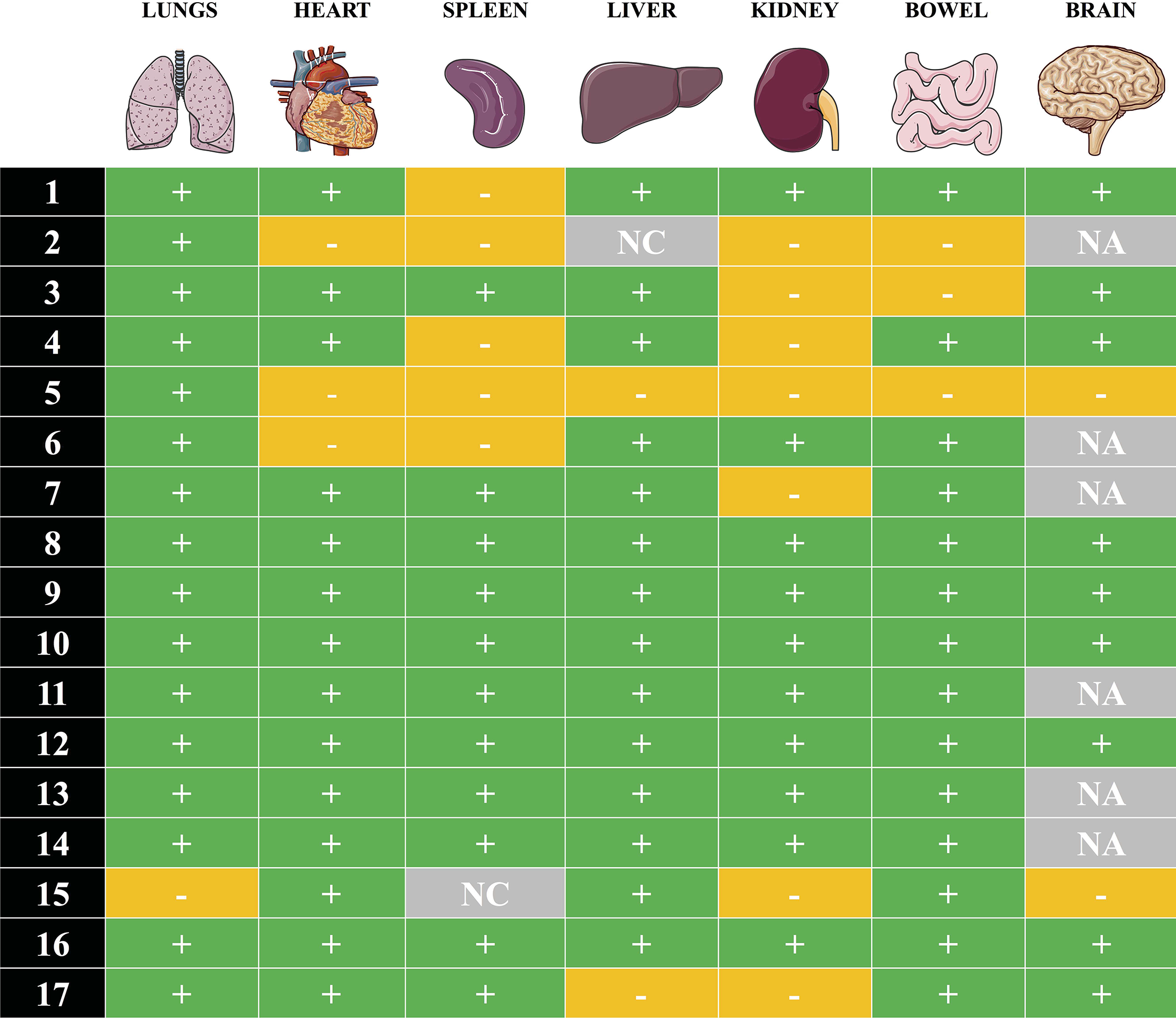
Detection of SARS-CoV-2 by immunohistochemistry (IHC) in FFPE *post-mortem* lung samples of 17 patients. Semi-quantitative evaluation: “-“negative result; “+”: scattered positive cells (between 1 and <5 positive cells/whole slide); “++”: positive isolated cells (> 5 cells/whole slide, but no foci); “+++”: foci of positive cells (more than 10 positive cells in one 200x field); NA = not available.

### SARS-CoV-2 detection by RT-PCR

SARS-CoV-2 RNA was detected in at least one organ from every patient (Figure 4). In the lung, RT-PCR was positive in 16 patients, with threshold cycle (Ct) values varying from 16.02 to 33.03. Viral RNA was also detected in the heart (n=14), the liver (n=14), the bowel (n=14), the spleen (n=11), and the kidney (n=10) as well as in 9 of the 11 cerebral samples. Ct values for non-pulmonary organs ranged from 28.67 to 35.11. Eight patients had positive RT-PCR in all tested organs.

**Figure 4:**
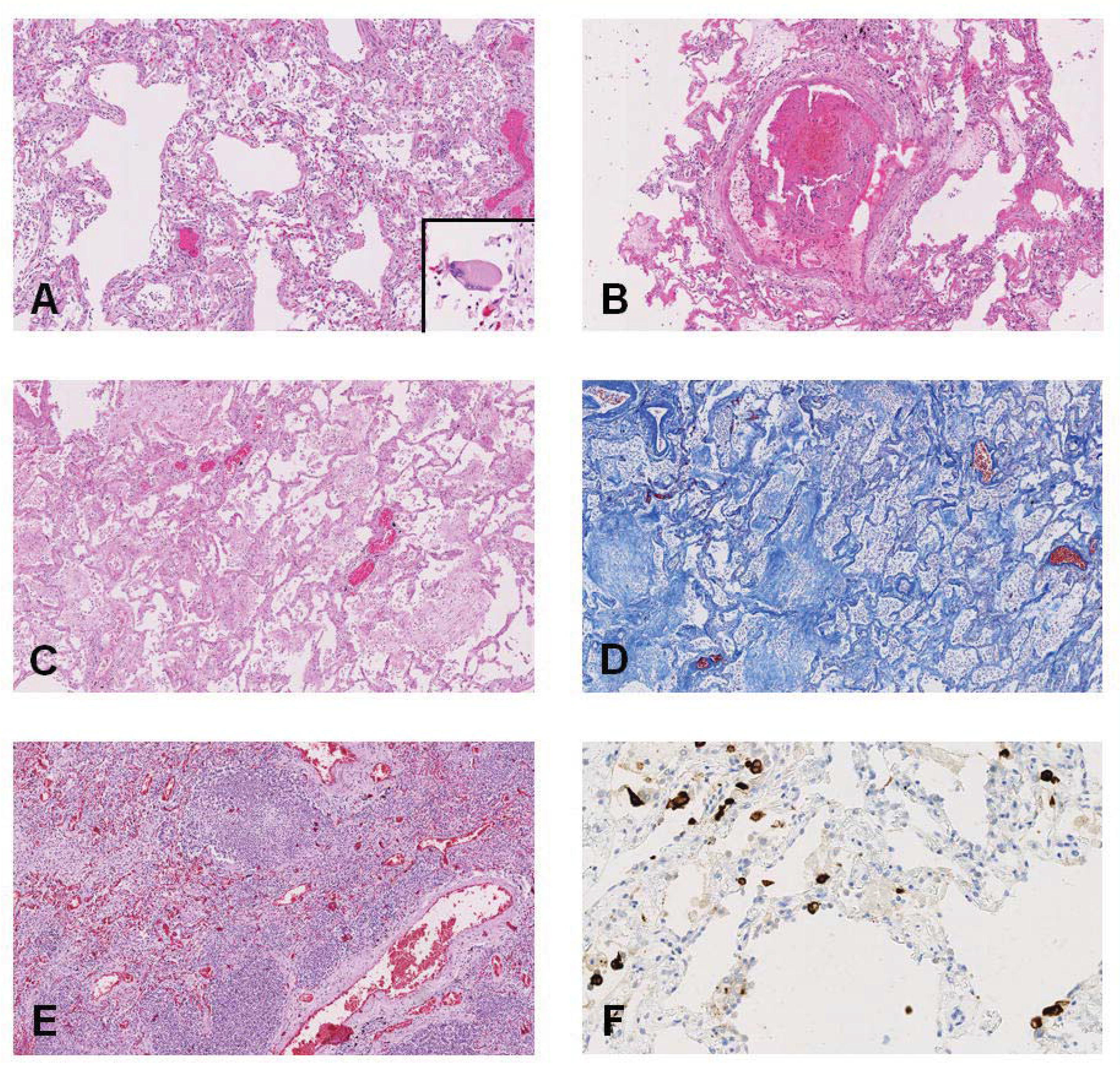
Molecular detection of SARS-Cov-2 RNA in *post-mortem* samples. Detection of SARS-CoV-2 by reverse transcription real-time polymerase chain reaction (RT-PCR) in FFPE *post-mortem* tissues of 17 patients. “+” positive result; “negative result; “NA” tissue not available, NC: non-informative test result (due to low quality RNA).

## Discussion

This *post-mortem* study showed several histopathological abnormalities in COVID-19 non-survivors; however, none of the findings was specific for direct viral injury. Moreover, the distribution of the infected cells was heterogeneous within the lung parenchyma. Finally, using RT-PCR, the virus could be detected in all examined organs.

The diagnosis of SARS-CoV-2-related organ injury is challenging; *post-mortem* histological findings were heterogeneous and often associated with chronic underlying diseases. In a previous autopsy study in COVID-19 patients (3), the authors reported that DAD associated with viral pneumonia was almost impossible to distinguish from that caused by bacterial pneumonia. No obvious intranuclear or intracytoplasmic viral inclusions were identified in another report (6). Desquamation of pneumocytes and hyaline membrane formation are frequently described in ARDS of many different causes, especially in early-phase ARDS (13). The presence of multinucleated cells with nuclear atypia is used to diagnose herpes virus infection in daily practice. As in previous reports (6, 14), we also observed the presence of multinucleated cells within lung alveoli in three patients; however, the significance of multinucleated cells is unclear and may not be specific of SARS-CoV-2 infection (15). Finally, some of the microscopic features of these patients are compatible with organ changes secondary to shock or systemic inflammation and no histological finding could be specifically ascribed to SARS-CoV-2.

In the absence of typical *post-mortem* viral features, our results show that RT-PCR is feasible on FFPE blocks and could be used in *post-mortem* analyses to identify the presence of SARS-CoV-2 in multiple organs and to understand the spread of the virus within the human body. The discordant RT-PCR and IHC results for detection of SARS-CoV2 in the lungs may be explained by the different sensitivity of these assays, which was higher for the RT-PCR, whereas low-level viral replication might not be detected by IHC. Moreover, IHC was based on the only available antibodies, which are targeted against SARS-CoV. New antibodies against SARS-CoV-2 need to be developed to improve the accuracy of IHC in the analysis of tissue samples from suspected or confirmed COVID-19 patients.

Most of the previous *post-mortem* studies in COVID-19 patients were conducted using needle biopsies and were therefore rather limited in terms of sampling; our complete autopsy analysis identified considerable heterogeneity of SARS-CoV-2 spread through the human body, and provides a more accurate description of macroscopic and microscopic organ alterations. As for previous coronavirus diseases (16, 17), the lungs are the most affected organs in COVID-19. However, DAD findings were highly heterogeneous, including both early-onset and additional late lesions. This finding could be explained by the heterogeneity of the pulmonary injury, including compliant lungs in the early phase and a more dense and non-recruitable lung in the late phase (18). As some patients died outside the ICU without undergoing mechanical ventilation, we could not estimate lung compliance before death. The heterogeneity could also reflect different treatments (e.g. fluid administration or corticosteroids) or different complications; as an example, half of the patients had concomitant acute pneumonia and it is difficult to conclude whether the DAD reflected the natural time-course of the viral disease or was secondary to superimposed complications, such as nosocomial infections. In a recent report, needle *post-mortem* biopsies suggested that COVID-19 is not associated with DAD but rather with an acute fibrinous and organizing pneumonia (AFOP), consequently requiring corticoid treatment (19). A diagnosis of AFOP is based on the absence of hyaline membranes and the presence of alveolar fibrin balls; however, hyaline membranes are heterogeneously distributed in the lung parenchyma with DAD and complete lung analysis, not just biopsies, are necessary to exclude their presence. Moreover, AFOP may be a fibrinous variant of DAD (20). The limitation of lung biopsy was also shown in another study, in which only 50% of lung samples were positive for SARS-CoV-2 using RT-PCR (21), when compared to almost 100% in our series. In addition, we did not find specific “endothelitis” as previously reported in a small case series (4). Considering the heterogeneity of *post-mortem* COVID-19 associated lesions, molecular and IHC assessment are mandatory in the histological analysis of COVID-19 tissue samples.

Patients with COVID-19 often have altered coagulation and a prothrombotic status, with the possible development of acute pulmonary embolism (PE) (22). In our study, three patients had PE, already diagnosed before death. Four patients had pulmonary infarction. In a previous study, acute PE was considered as the main cause of death in four patients (3); however, the inclusion of patients who died before hospital admission and the lack of specific thromboprophylaxis during the hospital stay may account for the differences in the severity of PE when compared to our study. Although we frequently observed the presence of microthrombi in the lung parenchyma, this feature is also reported in other forms of ARDS, regardless of etiology (13, 23). As such, whether diffuse pulmonary thrombosis is a main contributor of the fatal course of severe hypoxemia in COVID-19 patients remains to be further studied.

We did not observe specific viral organ injury, such as myocarditis, hepatitis or encephalitis. The cases of “acute cardiac injury” reported in COVID-19 clinical studies (24), do not necessarily translate into myocarditis or acute myocardial ischemia (only two had acute myocardial ischemia), similar to data reported in septic patients (i.e. elevated troponin without overt cardiac ischemia) (25). However, using RT-PCR, we found the virus in almost all the examined organs; this suggests that the virus can link to most cells, probably via the ACE2 receptor, which is ubiquitous, but may not directly cause organ injury. As extra-pulmonary direct viral injury (e.g. encephalitis, hepatitis, or myocarditis) has only been reported in very few cases, we suggest that SARS-CoV-2 infection may be just the trigger for an overwhelming host response, which could secondarily result in COVID-19-associated organ dysfunction. As RT-PCR might just detect residual viral genome, it remains unclear whether this represents active viral replication into the tissues or previous cellular infection, without clinically relevant significance (26).

This study has several limitations. First, the sample size was relatively small, and autopsies were only carried out from 72 to 96 hours after death. This delay did not allow us to properly analyze the gastrointestinal tract and kidneys, which showed signs of autolysis; in particular, acute tubular injury in the proximal tubules was indistinguishable from autolysis. Second, we could not determine the time-course and/or sequence of organ spread of the virus and no specific hypothesis regarding how SARS-CoV-2 spread (e.g. hematogenously) could be identified. Third, the time to death differed from patient to patient as did the course of the disease and treatments received, which limits a precise clinical-pathological correlation of histological findings related to COVID-19. Finally, we did not evaluate specific mechanisms involved in the pathogenesis of organ injury.

## Conclusion

These results underline the heterogeneity of organ injuries during COVID-19 disease and the absence of specific SARS-CoV-2 lesions. Using RT-PCR, SARS-CoV-2 could be detected in all organs, even those without evident microscopic lesions.

## Data Availability

The data that support the findings of this study are available from the corresponding author on reasonable request. Participant data without names and identifiers will be made available after approval from the corresponding author and local Ethics Committee. The research team will provide an email address for communication once the data are approved to be shared with others. The proposal with detailed description of study objectives and statistical analysis plan will be needed for evaluation of the reasonability to request for our data. Additional materials may also be required during the process.

**List of abbreviations**
ACE2: angiotensin converting enzyme 2
AFOP: acute fibrinous and organizing pneumonia
AKI: acute kidney injury
ARDS: Acute respiratory distress syndrome
COVID-19: coronavirus disease 2019
Ct: threshold cycle
DAD: diffuse alveolar damage
FFPE: Formalin-fixed paraffin-embedded
H&E: hematoxylin and eosin
IHC: immunohistochemistry
IQRs: interquartile ranges
MERS-CoV: Middle East respiratory syndrome coronavirus
PAS: periodic acid-Schiff
PE: pulmonary embolism
RT-PCR: real-time reverse-transcription polymerase chain reaction
SARS-CoV: severe acute respiratory syndrome coronavirus

## Declarations

### Ethics approval

The study protocol was approved by the local ethics committee (Erasme Hospital P2020/218). The ethical committee has waived the need for written informed consent.

### Consent for publication

Not applicable

### Authors’ contributions

IS had the idea for and designed the study and had full access to all the data in the study and takes responsibility for the integrity of the data and the accuracy of the data analysis.

IS, FT, JLV, and CD drafted the paper.

MR, CV, LL, PL, MLR, CM, ALT, JCG, LP, RDM, SD, SR, ND, LP, OD collected the data.

MR, ND, RDM did the analysis, and all authors critically revised the manuscript for important intellectual content and gave final approval for the version to be published. All authors agree to be accountable for all aspects of the work in ensuring that questions related to the accuracy or integrity of any part of the work are appropriately investigated and resolved.

## Acknowledgments

The authors thank Nathalie Lijsen, Christophe Valleys, Barbara Alexiou, Dominique Penninck, Nicole Haye and Audrey Verrellen for technical and logistic supports, Prof Frédéric Schuind for neurosurgical procedure, Egor Zindy (DIAPath, ULB) for proofreading the paper and Dr Marie-Paule Van Craynest for trainees’ supervision.

## Additional files

**Additional file 1:**

Critical care-autopsy-Covid-Additional file 1.doc

Additional material

Procedure to obtain brain samples

**Additional file 2:**

Critical care-autopsy-Covid-Additional file 2.doc

Additional table S1

Laboratory findings on the day of admission

**Additional file 3:**

Critical care-autopsy-Covid-Additional file 3.doc

Additional table S2

Detailed histological findings in all patients

## References

1. Guan WJ, Ni ZY, Hu Y, Liang WH, Ou CQ, He JX, et al. Clinical Characteristics of Coronavirus Disease 2019 in China. N Engl J Med. 2020;382:1708–20

2. Hoffmann M, Kleine-Weber H, Schroeder S, Krüger N, Herrler T, Erichsen S, et al. SARS-CoV-2 cell entry depends on ACE2 and TMPRSS2 and is blocked by a clinically proven protease inhibitor. Cell. 2020;181:1–10.

3. Wichmann D, Sperhake JP, Lütgehetmann M, Steurer S, Edler C, Heinemann A, et al. Autopsy Findings and Venous Thromboembolism in Patients With COVID-19: A Prospective Cohort Study. Ann Intern Med. 2020. Epub ahead of print. doi: 10.7326/M20-2003

4. Varga Z, Flammer AJ, Steiger P, Haberecker M, Andermatt R, Zinkernagel AS, et al. Endothelial cell infection and endotheliitis in COVID-19. Lancet. 2020; 395: 1417–8.

5. Chen T, Wu D, Chen H, Yan W, Yang D, Chen G, et al. Clinical characteristics of 113 deceased patients with coronavirus disease 2019: retrospective study. BMJ. 2020. Epub ahead of print. doi: 10.1136/bmj.m1091

6. Xu Z, Shi L, Wang Y, Zhang J, Huang L, Zhang C, et al. Pathological findings of COVID-19 associated with acute respiratory distress syndrome. Lancet Respir Med. 2020;8:420–2.

7. Barton LM, Duval EJ, Stroberg E, Ghosh S, Mukhopadhyay S. COVID-19 autopsies, Oklahoma, USA. Am J Clin Pathol. 2020;153:725–33.

8. ARDS Definition Task Force, Ranieri VM, Rubenfeld GD, Thompson BT, Ferguson ND, Caldwell E, Fan E, et al. Acute respiratory distress syndrome: the Berlin Definition. JAMA. 2012;307:2526–33.

9. Kellum JA, Lameire N; KDIGO AKI Guideline Work Group. Diagnosis, evaluation, and management of acute kidney injury: a KDIGO summary (Part 1). Crit Care. 2013;17:204.

10. Procédure pour les hôpitaux: prise en charge d’un patient possible ou confirmé COVID-19. https://epidemio.wiv-isp.be/ID/Documents/Covid19/COVID-19)_procedure_deaths_FR.pdf. Accessed 03/16/20.

11. D’Haene N, Meléndez B, Blanchard O, De Nève N, Lebrun L, Van Campenhout C, et al. Design and Validation of a Gene-Targeted, Next-Generation Sequencing Panel for Routine Diagnosis in Gliomas. Cancers (Basel). 2019;11:773.

12. Corman VM, Landt O, Kaiser M, Molenkamp R, Meijer A, Chu DK, et al. Detection of 2019 novel coronavirus (2019-nCoV) by real-time RT-PCR. Euro Surveill. 2020;25: 2000045.

13. de Hemptinne Q, Remmelink M, Brimioulle S, Salmon I, Vincent JL. ARDS: a clinicopathological confrontation. Chest. 2009;135: 944–9.

14. Menter T, Haslbauer JD, Nienhold R, Savic S, Hopfer H, Deigendesch N, et al. Post-mortem examination of COVID19 patients reveals diffuse alveolar damage with severe capillary congestion and variegated findings of lungs and other organs suggesting vascular dysfunction. Histopathology. 2020. Epub ahead of print. doi: 10.1111/his.14134.

15. Franks TJ, Chong PY, Chui P, Galvin JR, Lourens RM, Reid AH, et al. Lung pathology of severe acute respiratory syndrome (SARS): a study of 8 autopsy cases from Singapore. Hum Pathol. 2003;34:743–8.

16. Nicholls JM, Poon LL, Lee KC, Ng WF, Lai ST, Leung CY, et al. Lung pathology of fatal severe acute respiratory syndrome. Lancet. 2003;361:1773–8.

17. Hwang, D., Chamberlain, D., Poutanen, S. Low DE, Asa SL, Butany J. Pulmonary pathology of severe acute respiratory syndrome in Toronto. Mod Pathol. 2005;18:1–10.

18. Gattinoni L, Chiumello D, Rossi S. COVID-19 pneumonia: ARDS or not? Crit Care. 2020;24:154.

19. Copin MC, Parmentier E, Duburcq, T Poissy J, Mathieu D and The Lille COVID-19 ICU and Anatomopathology Group. Time to consider histologic pattern of lung injury to treat critically ill patients with COVID-19 infection. Intensive Care Med. 2020 Apr 23: 1–3.

20. Santos C, Oliveira RC, Serra P, Baptista JP, Sousa E, Casanova P, et al. Pathophysiology of acute fibrinous and organizing pneumonia - Clinical and morphological spectra. Pathophysiology. 2019; 26: 213–7.

21. Wang D, Hu B, Hu C, Zhu F, Liu X, Zhang J, et al. Clinical Characteristics of 138 Hospitalized Patients With 2019 Novel Coronavirus-Infected Pneumonia in Wuhan, China. JAMA. 2020. Epub ahead of print. doi: 10.1001/jama.2020.1585.

22. Llitjos JF, Leclerc M, Chochois C, Monsallier JM, Ramakers M, Auvray M, et al. High incidence of venous thromboembolic events in anticoagulated severe COVID-19 patients. J Thromb Haemost. 2020. Epub ahead of print. doi: 10.1111/jth.14869.

23. Chang JC. Acute Respiratory Distress Syndrome as an Organ Phenotype of Vascular Microthrombotic Disease: Based on Hemostatic Theory and Endothelial Molecular Pathogenesis. Clin Appl Thromb Hemost. 2019;25:1076029619887437.

24. Shi S, Qin M, Shen B, Cai Y, Liu T, Yang F, et al. Association of cardiac injury with mortality in hospitalized patients with COVID-19 in Wuhan, China. JAMA Cardiol. 2020. Epub ahead of print. doi: 10.1001/jamacardio.2020.0950.

25. Smeding L, Plötz FB, Groeneveld AB, Kneyber MC. Structural changes of the heart during severe sepsis or septic shock. Shock. 2012;37:449–56.

26. Wölfel R, Corman VM, Guggemos W, Seilmaier M, Zange S, Müller MA, et al. Virological assessment of hospitalized patients with COVID-2019. Nature. 2020. Epub ahead of print. doi: 10.1038/s41586-020-2196-x.

